# Long-term risk of Stroke after Acute Coronary Syndrome. The ABC-10* Study on Heart Disease

**DOI:** 10.1101/2024.05.29.24308175

**Authors:** Heba T. Mahmoud, Rocco Cordiano, David Merotto, Mattia Ludovico Dario, Fiorella Cavuto, Giuseppe Berton

## Abstract

**Background:** Previous studies link myocardial infarction to increased stroke risk. This long-term prospective study examines stroke incidence and outcomes in acute coronary syndrome (ACS) patients, identifying risk factors and geographic disparities.

**Methods:** We enrolled 535 ACS patients admitted to hospitals across three provinces in the Veneto region of Italy. Patients’ residences were classified into three urban and three rural areas in each province. Patients were followed prospectively for 24 years or until death.

**Results:** All patients, except for three, completed the follow-up, totaling 6151 person-years. During follow-up, 84 patients experienced a stroke, with 85% being ischemic and 15% hemorrhagic, proving fatal in 43 cases. The stroke incidence rate was 14/1000 person-years. Multivariable Cox regression analysis identified older age (HR 1.84; 95% CI 1.30-2.60), atrial fibrillation (AF) (HR 2.64; 95% CI 1.49-4.67), and a higher albumin-to-creatinine ratio (HR 1.38; 95% CI 1.04-1.83) as independent predictors of overall stroke risk, while higher eGFR (HR 0.71; 95% CI 0.53-0.95) was independently associated with a lower risk.

A sub-analysis revealed older age (HR 2.67; 95% CI 1.60-4.45) and AF (HR 2.95; 95% CI 1.38-6.32) as independent predictors of fatal stroke. Unexpectedly, we observed a higher fatal stroke risk in urban areas (HR 1.89; 95% CI 1.03-3.48) and southern provinces (HR 1.71; 95% CI 1.15-2.53).

**Conclusion:** The ABC study identified several baseline clinical predictors associated with higher stroke risk long after ACS. A geographical association with the risk of fatal stroke was also observed, underscoring the importance of considering both individual clinical predictors and broader geographic factors in stroke prevention policies.

## INTRODUCTION

Globally, ischemic heart disease remains the number one cause of death, responsible for 16% of the world’s total deaths, while cerebral stroke is the second leading cause of death, responsible for approximately 11% of total deaths and the number one cause of disability. Both are the top two causes of death in Italy for both men and women [1].

Previous studies have documented an increased risk of stroke, mainly ischemic stroke (IS), in patients who survived myocardial infarction (MI), with a 1-year risk ranging from 2% to 4% [2–4]. Coronary heart disease and some subtypes of IS are linked by inflammation and the development of atherosclerosis, and share several risk factors including age, hypertension, dyslipidemia, smoking, and diabetes. Additionally, MI can itself be a risk factor for stroke through mechanisms such as emboli, either during revascularization or from atrial fibrillation (AF) in association with acute MI or from blood stasis in a poorly functioning left ventricle [3]. However, most of these reports were based on retrospective analysis and were limited to short follow-up duration. Moreover, relatively few studies have examined the incidence and long-term outcomes of stroke in patients with ACS without ST-segment elevation [4–6], and to our knowledge, there are no reports about the geographic differences in stroke risk in this specific population.

In the present long-term prospective study, we assessed the incidence and clinical outcome of stroke in an unselected sample of patients discharged alive after an index hospitalization with ACS and followed for 24 years. An additional study goal was to identify risk factors for stroke in these high-risk patients. We also investigated the existence of disparities in stroke risk across geographic areas within the Veneto region of Italy.

## METHODS

### Patients

The ABC Study on Heart Disease (https://www.abcstudy.foundation/) is an ongoing prospective study designed to represent, as closely as possible, an unbiased population of patients with ACS. Specifically, the study includes Caucasian patients who were admitted to the intensive care units of three general hospitals in Italy’s Veneto region between June 1995 and January 1998 with definite diagnosis of ACS, including ST-elevation myocardial infarction (STEMI), non-ST elevation myocardial infarction (NSTEMI), or unstable angina pectoris (UAP). The study originally aimed to monitor the long-term natural history of these patients, both non-fatal and fatal events, and causes of death. An additional aim of the study was to investigate the prognostic value of multiple baseline clinical variables. The diagnosis of ACS was based on the presence of at least two of the following criteria: typical changes in serum enzymes, such as total creatine kinase (CK) and creatine kinase MB (CK-MB), typical electrocardiogram changes (i.e., localized ST-T changes and/or pathological Q waves in at least two contiguous leads), and central chest pain that lasted more than 30 minutes [7].

Of the 741 consecutive unselected patients deemed eligible upon admission, the study excluded 84 patients due to diagnoses other than ACS, 23 with missing baseline data and 54 patients due to residing outside the Veneto region. Forty-five of the remaining 580 patients died during the index hospitalization, leaving 535 patients in the post-discharge follow-up **(Figure 1).** Each patient was assigned an anonymous code, and the baseline or follow-up database did not include any personal data or identifiers. The study protocol adhered to the Declaration of Helsinki and received approval from the ethics committee of each hospital. All enrolled patients provided written informed consent.

**Figure 1.**
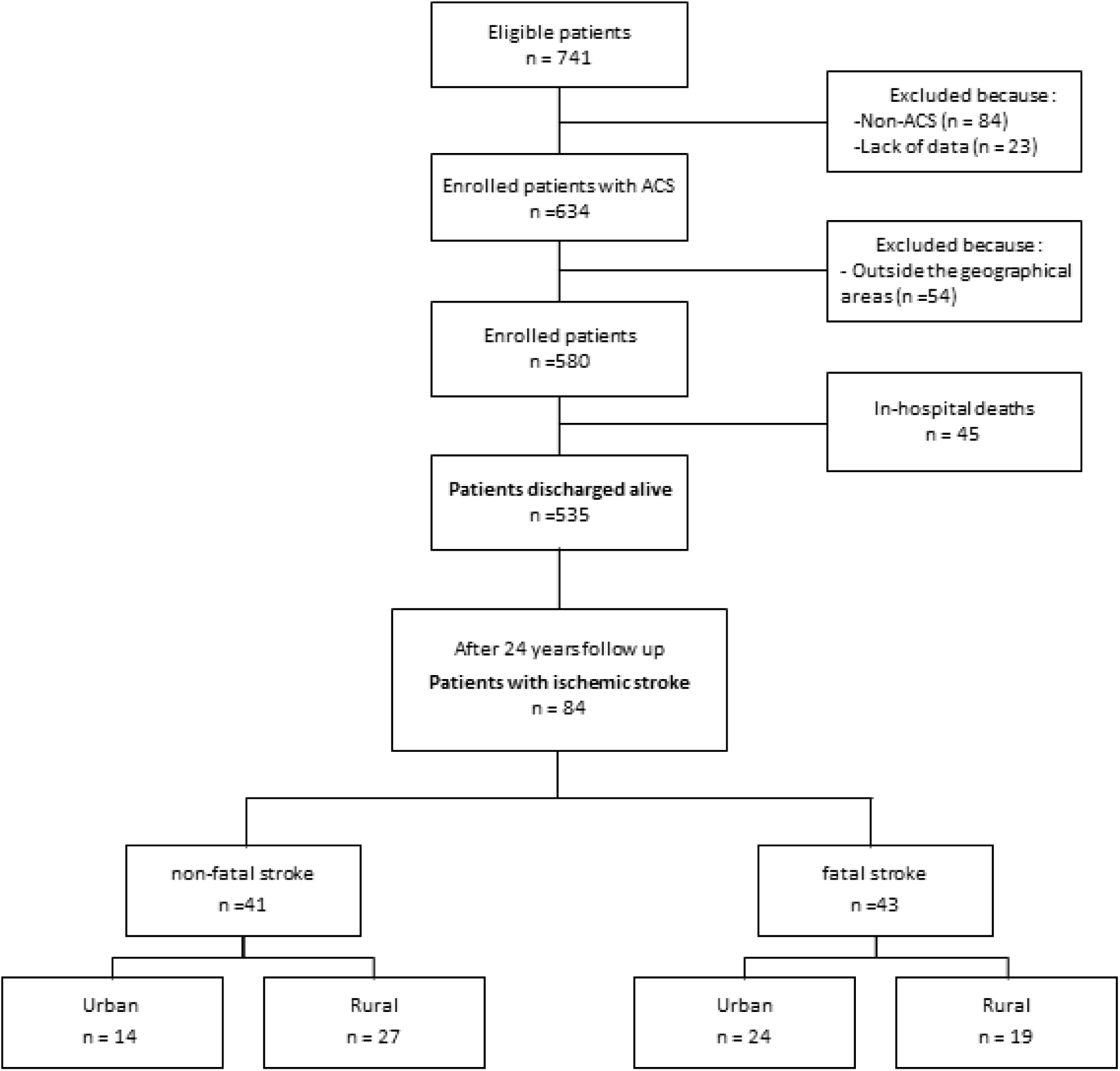
Flow diagram of the study population and progress during follow-up. **ACS:** acute coronary syndrome.

### Urban-rural classification

As previously described in detail [8], the ABC study includes patients admitted to hospitals in the following cities in Italy’s Veneto region: Conegliano-Vittorio Veneto in Treviso province (northern), Bassano in Vicenza province (central), and Adria-Cavarzere in Rovigo province (southern). Residency in each province was categorized as urban or rural using the Rural Development Programme (Programma di Sviluppo Rurale [PSR] 2014-2020) classification of municipalities [9]. The total population of the study area is 586,976, with 24% residing in urban areas and 76% in rural areas [9, 10].

### Measurements and follow-up

A thorough medical history was collected from the patient’s medical records and patient interviews at enrolment. All the analyzed baseline clinical and laboratory data were obtained during the first 7 days of hospitalization in the intensive coronary care unit as previously described in detail [11–13].

Clinical check-ups were done for each patient 1, 3, 5, 7, 10, 12, 15, 17, 20, 22 and 24 years after enrolment. At each recruitment hospital, two cardiologists were responsible for monitoring the cohort of patients throughout the follow-up.

For the present analysis, the pre-specified primary endpoint was the occurrence of cerebral stroke which was defined by the World Health Organization (WHO) criteria: a focal neurological disorder with a rapid onset that persists for at least 24h or until death, which includes ischemic stroke (IS) and hemorrhagic stroke (HS) (intracerebral and subarachnoid hemorrhage) [14]. Data were obtained from scheduled examinations, public administrations, hospital records, family doctors, post-mortem examinations, and death certificates. The medications received during the index hospitalization and follow-up treatments were also recorded. All post-enrolment data were recorded prospectively according to the ABC Study on Heart Disease protocol [12]. Baseline data and follow-up data were recorded in two different datasheets that were merged after the completion of 24 years of follow-up.

### Statistical analysis

Unpaired Student’s t-test and Pearson chi-square (χ^2^) test were used for measured and categorical variables, respectively. Log transformations were used to correct positive-skewed distributions, as appropriate. If a patient dropped out before 24 years of follow-up, her/his data were censored at that time.

In survival analysis, continuous variables were analyzed as terciles of increasing values. Survival curves were constructed using cumulative incidence as a function of incident stroke [15]. Stroke incidence rates with person-time denominators were calculated. Person-time at risk was accumulated from index admission for ACS until stroke onset, death, or end of follow-up, whichever came first. Cox proportional hazard regression analysis was performed to estimate the hazard ratios (HRs) and 95% confidence intervals (CIs) for stroke risk. Scaled Schoenfeld residuals were used to test the proportionality assumption with 95% CI.

Results were reported as medians and interquartile ranges for continuous variables and numbers and percentages for categorical variables. Unless otherwise indicated, two-tailed P values <0.05 were deemed significant. The statistical analyses were performed using STATA 18 (College Station, Texas, USA).

## RESULTS

### Patient Characteristics

Unless pre-empted by death, all enrolled patients completed the follow-up representing 6151 Person-years except for three patients for whom survival time was censored before 24 years: two withdrew consent and one moved overseas. The median age of the study population was 67 years, 70% were male, and 318 patients (59%) were residing in rural areas. ACS was distributed as follows: STEMI 62%, NSTEMI 21% and UAP 17%.

During the follow-up period, 84 patients (16%) experienced an acute stroke event, with 85% being ischemic and 15% hemorrhagic. The stroke proved fatal in 43 patients, Figure 1. Patients who experienced a stroke and those who did not, shared most of the baseline demographic and clinical characteristics as detailed in Table 1.

**Table 1.**
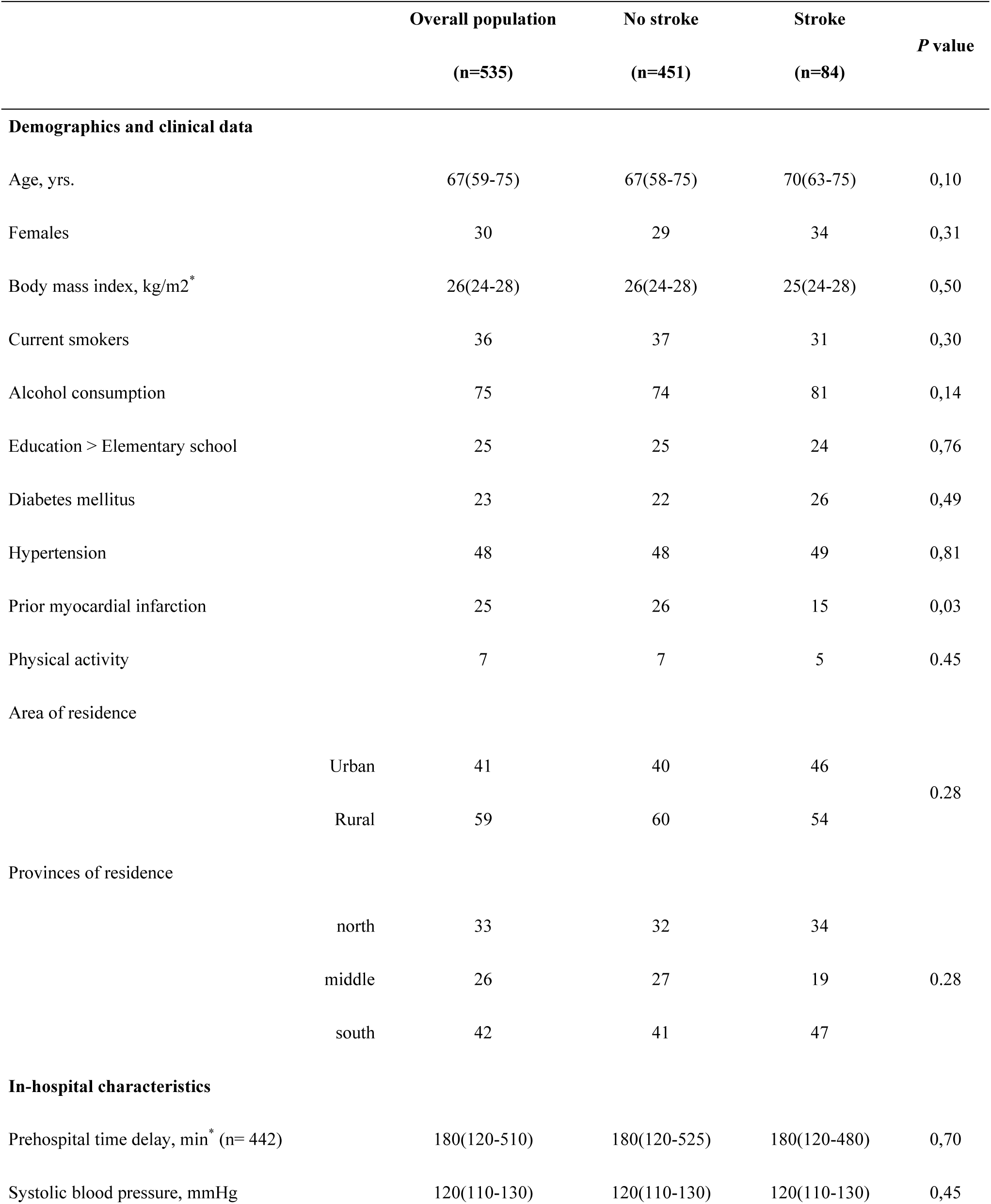
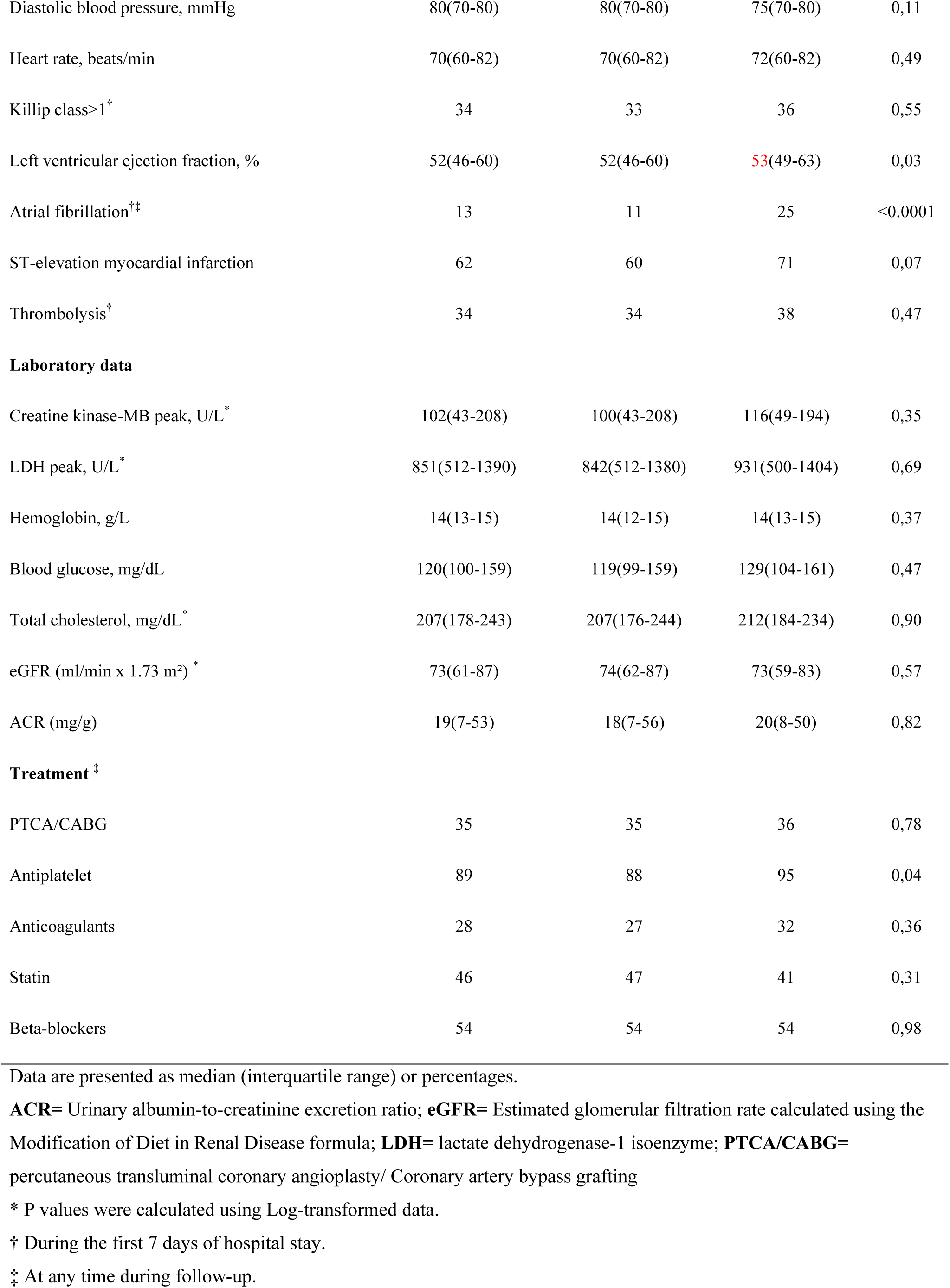
Baseline characteristics of ACS patients with and without stroke.

### Overall stroke risk during follow-up

The incidence rate (IR) of stroke during the entire span of the follow-up time was 14 per 1000 person-year. The median time from enrolment to first stroke diagnosis was 7 years (interquartile range (IQR): from 3 to 15 years). Stroke IR by different clinical variables are provided in Table 2. The urban/rural stroke IR per 1000 person-years was (17/15 in the north, 15/10 in the middle, and 17/11 in the southern province).

**Table 2:**
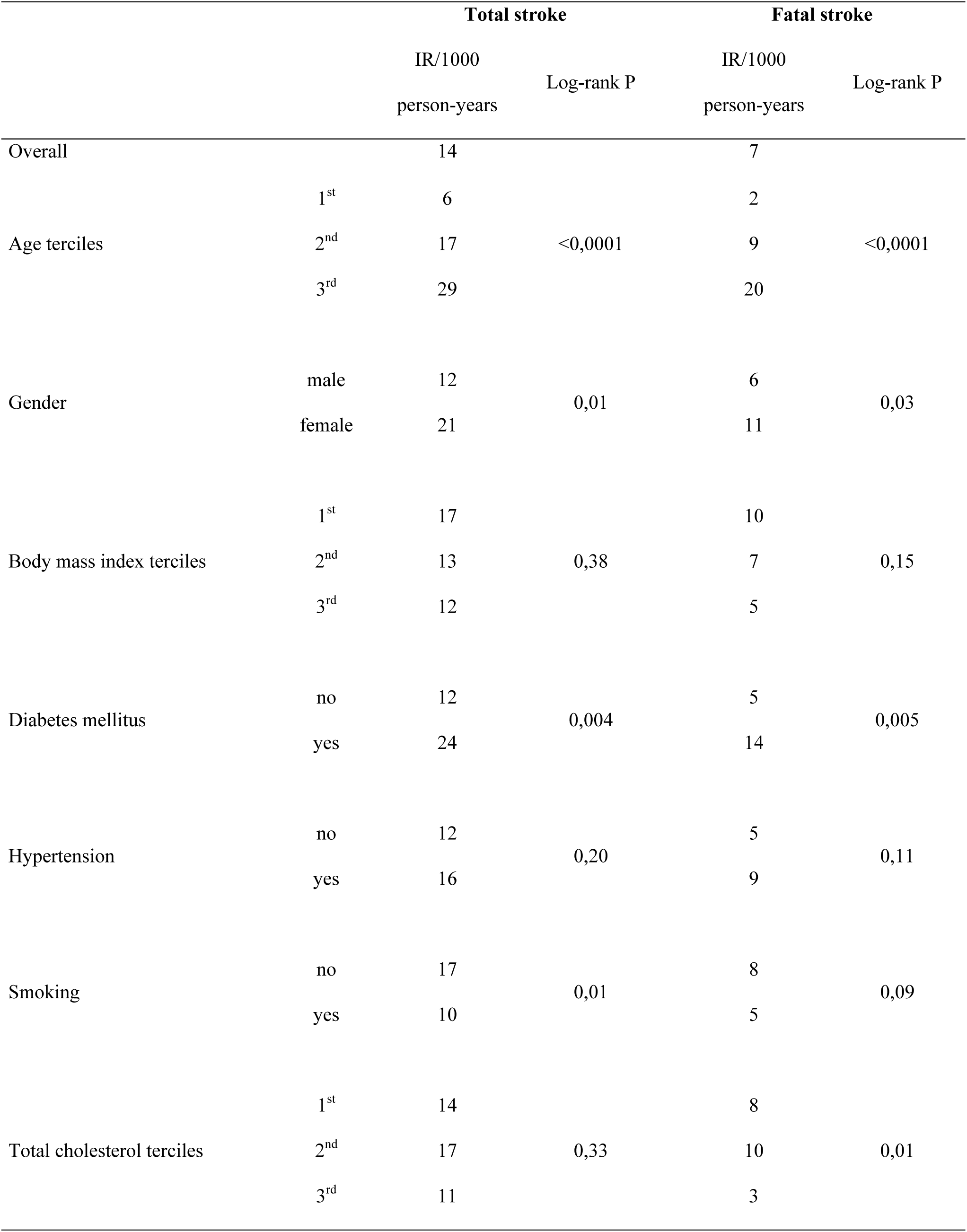
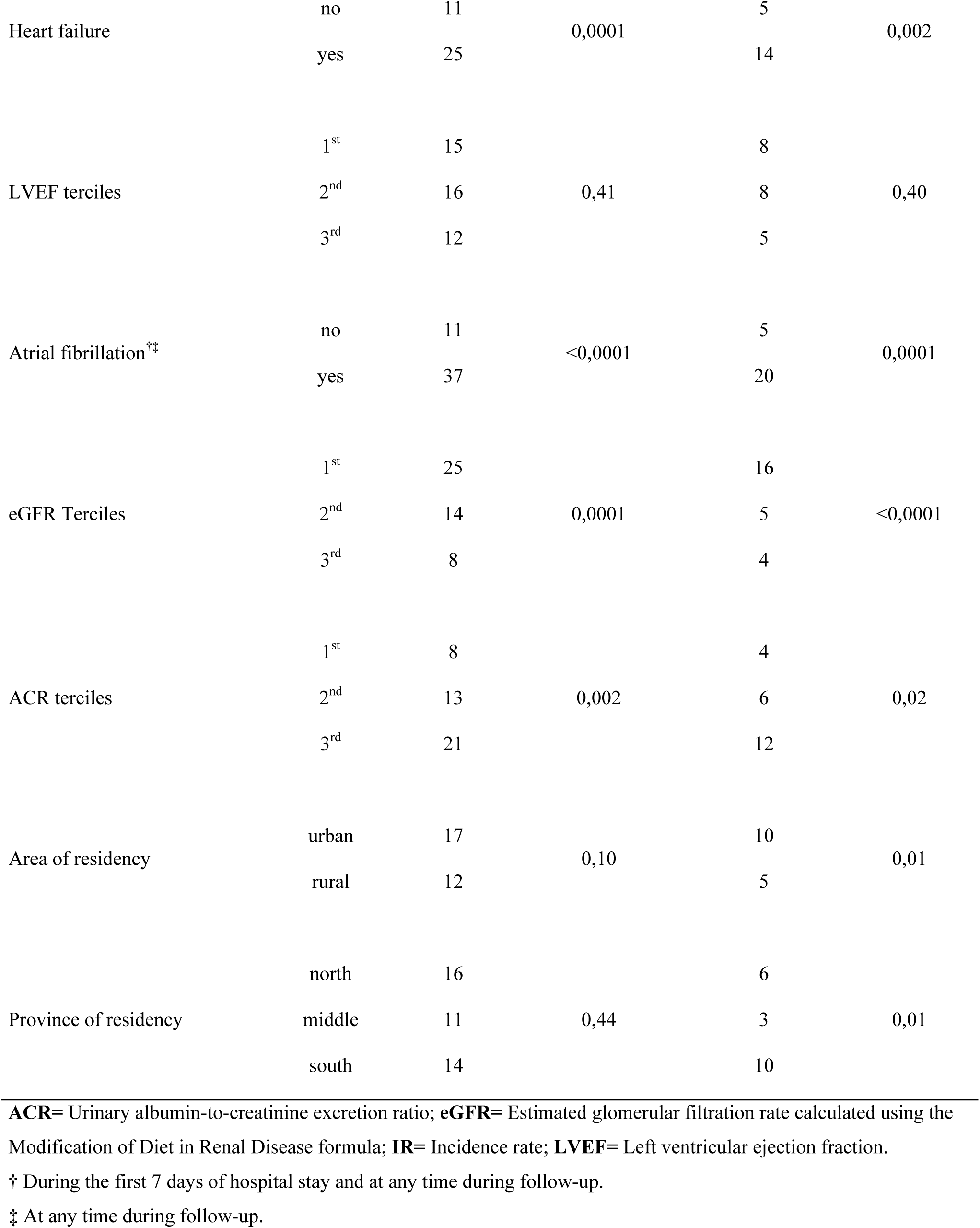
Incidence rate of total and fatal stroke 24 years after acute coronary syndrome.

Univariable Cox regression analyses of predictors of stroke within 24 years after ACS are shown in Table 3. Older age (HR 2.4; 95%CI 1.8-3.2), female gender (HR 1.8; 95%CI 1.1-2.8), baseline diabetes (HR 2.0; 95%CI 1.2-3.3), heart failure (HR 2.4; 95%CI 1.6-3.8), AF (HR 3.4; 95%CI 2.1-5.6) and higher albumin-to-creatinine ratio (ACR) (HR 1.7; 95%CI 1.2-2.3) were all associated with an increased risk of stroke. while higher eGFR, reperfusion, statin and beta-blockers treatment during follow-up were associated with a decreased risk; HR= (0.6(95%CI 0.4-0.7), 0.5(95%CI 0.3-0.8), and 0.4(95%CI 0.3-0.6), respectively).

**Table 3:** Uni- and multivariable analysis of predictors of overall stroke 24 years after ACS.

| Predictor of risk | Overall stroke risk |  | Fatal stroke |  |
| --- | --- | --- | --- | --- |
|  | Hazard ratio (95% CI) | p-value | Hazard ratio (95% CI) | p-value |
| **Univariable analysis** |  |  |  |  |
| Age terciles | 2,43 (1,81-3,27) | <0,0001 | 3,36 (2,17-5,20) | <0,0001 |
| Female gender | 1,79 (1,14-2,81) | 0,01 | 1,90 (1,02-3,54) | 0,04 |
| BMI terciles | 0,82 (0,63-1,08) | 0,16 | 0,69 (0,47-1,01) | 0,06 |
| Current smoking | 0,56 (0,35-0,89) | 0,01 | 0,58 (0,30-1,11) | 0,10 |
| Alcohol consumption | 1,40 (0,81-2,42) | 0,23 | 1,42 (0,66-3,07) | 0,37 |
| Education > Elementary school | 0,91 (0,68-1,21) | 0,50 | 0,77 (0,49-1,20) | 0,25 |
| Diabetes mellitus | 2,02 (1,24-3,30) | 0,005 | 2,46 (1,29-4,68) | 0,006 |
| Hypertension | 1,29 (0,84-1,99) | 0,24 | 1,61 (0,88-2,95) | 0,12 |
| Prior myocardial infarction | 0,87 (0,48-1,58) | 0,65 | 1,19 (0,57-2,48) | 0,65 |
| Physical activity | 0,51 (0,19-1,39) | 0,19 | 0,52 (0,13-2,15) | 0,37 |
| Residency area (rural-urban) | 1,39 (0,91-2,14) | 0,13 | 2,08 (1,14-3,80) | 0,02 |
| Residency provinces (north-middle-south) | 0,94 (0,73-1,20) | 0,61 | 1,47 (1,00-2,15) | 0,04 |
| Systolic blood pressure terciles | 1,00 (0,75-1,33) | 0,99 | 0,99 (0,66-1,48) | 0,96 |
| Diastolic blood pressure terciles | 0,88 (0,67-1,17) | 0,40 | 0,80 (0,53-1,20) | 0,28 |
| Heart rate terciles | 1,25 (0,94-1,67) | 0,13 | 1,46 (0,98-2,17) | 0,06 |
| Left ventricular ejection fraction | 0,89 (0,69-1,15) | 0,38 | 0,80 (0,55-1,14) | 0,22 |
| ST-elevation myocardial infarction | 1,38 (0,86-2,22) | 0,18 | 1,14 (0,60-2,17) | 0,68 |
| Killip class>1<sup>†</sup> | 2,44 (1,55-3,84) | <0,0001 | 2,56 (1,39-4,74) | 0,003 |
| Atrial fibrillation<sup>†‡</sup> | 3,40 (2,06-5,59) | <0,0001 | 3,59 (1,84-7,01) | <0,0001 |
| Thrombolysis<sup>†</sup> | 0,77 (0,50-1,21) | 0,26 | 0,59 (0,31-1,13) | 0,11 |
| Total cholesterol terciles | 0,89 (0,68-1,15) | 0,37 | 0,65 (0,44-0,95) | 0,03 |
| eGFR terciles | 0,55 (0,42-0,73) | <0,0001 | 0,46 (0,31-0,68) | <0,0001 |
| ACR terciles<sup>‡</sup> | 1,68 (1,24-2,27) | 0,001 | 1,72 (1,14-2,60) | 0,01 |
| PTCA/CABG<sup>‡</sup> | 0,50 (0,31-0,78) | 0,002 | 0,25 (0,12-0,54) | <0,0001 |
| Antiplatelet<sup>‡</sup> | 0,93 (0,34-2,57) | 0,90 | 1,10 (0,26-4,60) | 0,89 |
| Anticoagulants<sup>‡</sup> | 0,98 (0,62-1,57) | 0,94 | 0,53 (0,25-1,15) | 0,11 |
| Beta-blockers<sup>‡</sup> | 0,40 (0,26-0,62) | <0,0001 | 0,30 (0,16-0,56) | <0,0001 |
| **Multivariable analysis** |  |  |  |  |
| Age terciles | 1,84 (1,30-2,60) | 0,001 | 2,67 (1,60-4,45) | <0,0001 |
| Female gender | 1,01 (0,62-1,64) | 0,97 | 0,95 (0,49-1,84) | 0,88 |
| Atrial fibrillation<sup>†</sup> | 2,64 (1,49-4,67) | 0,001 | 2,95 (1,38-6,32) | 0,005 |
| eGFR terciles | 0,71 (0,53-0,95) | 0,02 | 0,66 (0,43-1,02) | 0,06 |
| ACR terciles | 1,38 (1,04-1,83) | 0,03 | 1,38 (0,93-2,06) | 0,11 |
| Area (rural-urban) | 1,25 (0,81-1,94) | 0,31 | 1,89 (1,03-3,48) | 0,04 |
| Provinces (north-middle-south) | 1,04 (0,80-1,36) | 0,76 | 1,71 (1,15-2,53) | 0,008 |

**BMI**= Body mass index; **eGFR**= Estimated glomerular filtration rate calculated using the Modification of Diet in Renal Disease formula; **ACR**= Urinary albumin-to-creatinine excretion ratio; **PTCA/CABG**= percutaneous transluminal coronary angioplasty/Coronary artery bypass grafting.
<sup>†</sup> During the first 7 days of hospital stay.
<sup>‡</sup> At any time during follow-up.

In multivariable analysis: older age, atrial fibrillation and higher ACR were independent predictors of the stroke, while higher eGFR was independently associated with a lower risk, Table 3 and Figure 2.

**Figure 2.**
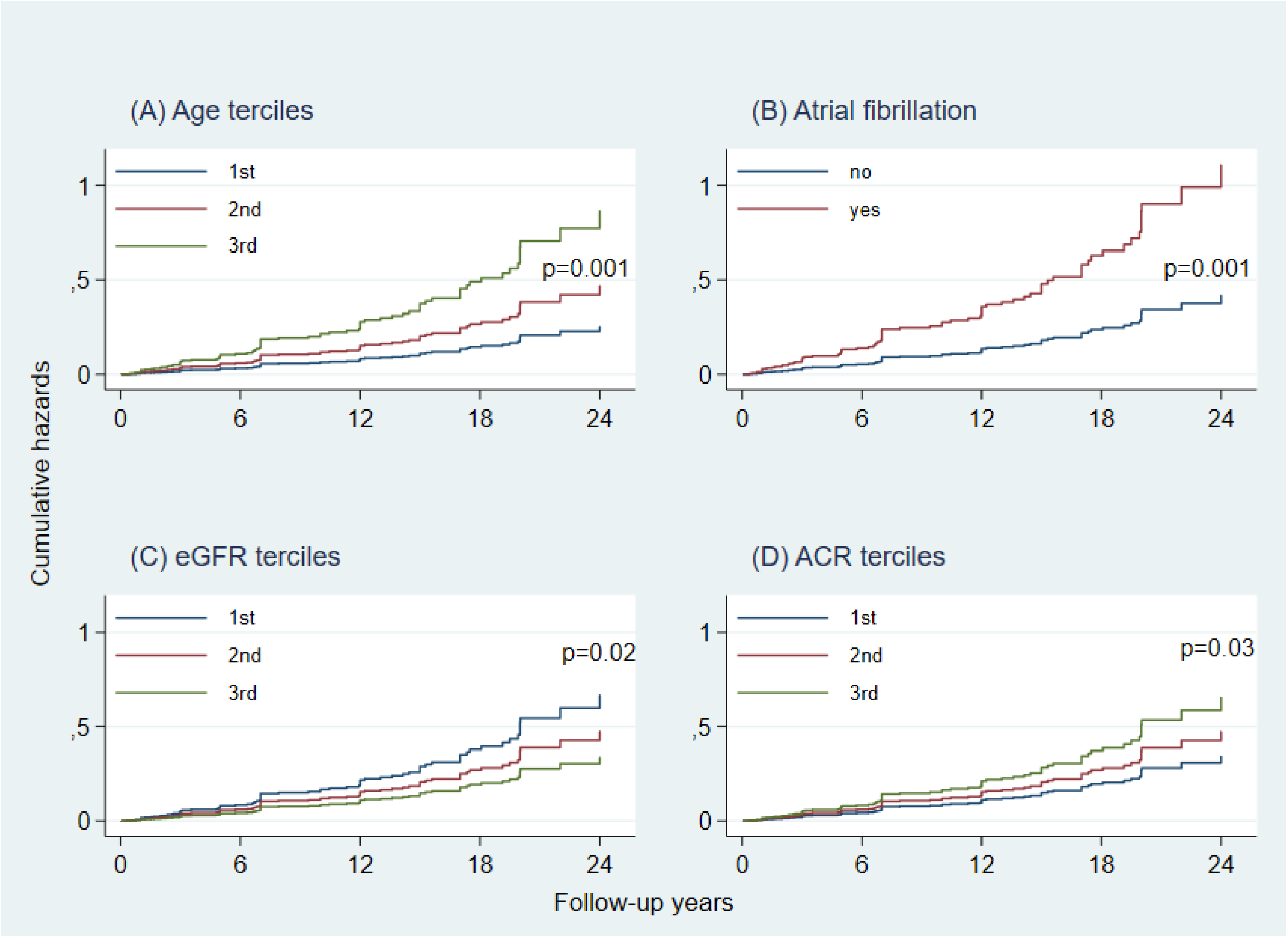
Fully adjusted cumulative hazard function of overall stroke by different clinical variables. **eGFR:** Estimated glomerular filtration rate calculated using the Modification of Diet in Renal Disease formula; **ACR:** Urinary albumin-to-creatinine excretion ratio.

### Risk of fatal stroke

A sub-analysis of the 43 patients who had a fatal stroke revealed an IR of 7 per 1000 person-years, Table 2. The median time from enrolment to fatal stroke was 6 years (IQR 3-13 years). The urban/rural incidence rate of fatal stroke per 1000 person-years was (7/5 in the north. 4/2 in the middle. and 15/7 in the southern province).

Univariable Cox regression analyses of predictors of fatal stroke within 24 years after ACS showed similar results as older age, female gender, baseline diabetes, heart failure, AF, and higher albumin-to-creatin ratio were all associated with an increased risk of fatal stroke, Table 3. Higher eGFR, reperfusion, and beta-blocker treatment during follow-up were associated with a decreased risk.

At the multivariable analysis, older age and AF were independent predictors of fatal stroke, Table 3.

Unexpectedly, we also observed an association between geographic areas of residence and the long-term risk of fatal stroke as the risk increased going from rural to urban areas (HR 2.1; 95%CI 1.1-3.8) and from north to middle and south provinces (HR 1.5; 95%CI 1.0-2.1) with the univariable Cox regression analysis. Results were kept true using multivariable Cox regression models, Table 3 and Figure 3.

**Figure 3.**
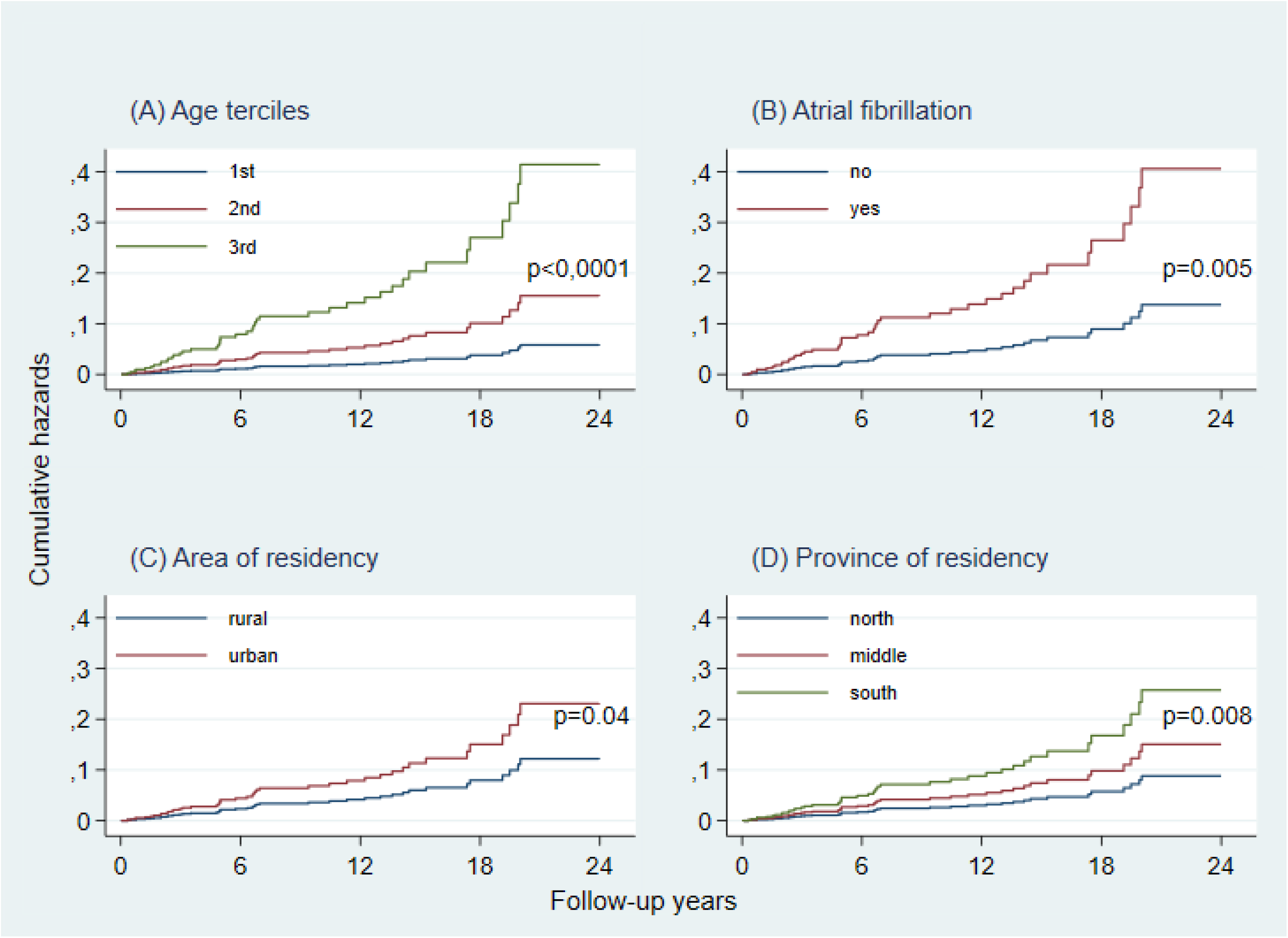
Fully adjusted cumulative hazard function of fatal strokes by different clinical variables.

### Ischemic and hemorrhagic strokes

In a further sub-analysis, we found that ischemic strokes predominated among the long-term events in this study, showing an IR of 12 per 1000 person-years, while hemorrhagic strokes exhibited a lower IR of 2 per 1000 person-years.

In the multivariable analysis, several independent predictors for IS, including older age (HR 2.4; 95%CI 1.6 - 3.5), AF (HR 2.0; 95%CI 1.0 - 3.7), and higher ACR (HR 1.5; 95%CI 1.1 - 2.0). On the other hand, for HS, only a higher eGFR (HR 0.2; 95%CI 0.1 – 0.5) was identified as a predictor for lower risk.

## DISCUSSION

Stroke represents a formidable challenge in healthcare irrespective of the underlying clinical condition due to its substantial impact on morbidity and mortality [16]. The ABC study offered crucial insights into the incidence and the enduring risk factors for stroke among patients with ACS. Specifically, it shed light on several baseline clinical predictors correlated with an increased risk of long-term stroke, such as older age, AF, and a higher ACR. In contrast, the study found that a higher eGFR was associated with a reduced risk of stroke in this particular population. These findings provide valuable guidance for healthcare professionals in identifying and managing stroke risk factors among ACS patients, ultimately contributing to improved patient outcomes and quality of care. Moreover, the study identified a geographical association with the risk of fatal stroke, highlighting the significance of understanding regional differences in stroke risk and implementing targeted preventive strategies.

Our study revealed an overall stroke incidence rate of 14 per 1000 person-years among patients with ACS with a cumulative incidence surpassing 15%, consistent with findings from prior investigations. For instance, Hurskainen et al. retrospectively documented a cumulative stroke incidence exceeding 10% during a 14-year follow-up of 8,049 ACS patients [6]. Long-term stroke is a critical outcome among patients with ACS, given its significantly high associated mortality. Notably, approximately half of the post-discharge strokes in our cohort resulted in fatalities, a finding also in line with previous research involving ACS patients [6, 17].

The mechanisms contributing to the elevated risk of stroke persisting for up to 25 years after ACS likely involve inflammation and shared risk factors for atherosclerosis. These factors contribute to mutual pathophysiological disarray underlying both diseases [6, 18].

The present study has identified several important risk factors for long-term stroke in patients with ACS. In particular, older age and AF were found to be independent predictors for both total and fatal stroke which is consistent with the findings of previous studies [17]. AF is an established risk factor for stroke, primarily due to its association with the formation of cardiac emboli. In the setting of AMI, AF can manifest either as a pre-existing condition or as a complication. Previous studies indicate an incidence ranging from 2-21% for AF complicating AMI [19, 20]. Consistent with our results, studies also showed that patients with AF following AMI had a significantly higher long-term risk for fatal and non-fatal stroke compared to those without AF regardless of their age, gender or interventional treatment [20, 21].

Our study integrates and advances existing literature on the association between eGFR, ACR, and stroke risk, demonstrating that patients with lower eGFR or higher ACR levels have an increased stroke risk [22, 23]. Notably, we found that higher eGFR predicts lower hemorrhagic stroke (HS) risk, consistent with findings from the Rotterdam study by Bos et al., which followed 4937 participants for an average of 10.2 years. In that study, decreased eGFR was a strong risk factor for hemorrhagic stroke, but not for ischemic stroke [24].

Our findings also corroborate the extensive body of evidence from epidemiological studies indicating that geographic differences in stroke mortality are genuine and not merely a result of reporting biases or data collection inconsistencies. Several factors, including socioeconomic status, lifestyle and dietary habits, genetic and biological characteristics, and environmental factors influence these disparities [25–27].

Given the potentially devastating consequences of stroke, it’s crucial to identify high-risk ACS patients promptly. This facilitates the implementation of targeted management strategies, such as aggressive risk factor modification, optimized medical therapy, and vigilant monitoring. These proactive approaches not only help reduce stroke incidence but also enhance overall patient outcomes while alleviating the strain on healthcare resources.

## LIMITATIONS

Our prospective study boasts several notable strengths. Chief among them is the exceptionally prolonged follow-up period, which spans an impressive 24 years and remarkably features minimal dropouts. To our knowledge, our research stands as the pioneering endeavor in examining the prognostic insights offered by baseline clinical variables over such an extended timeframe following ACS. Furthermore, the inclusion of comprehensive baseline characteristics, ongoing medical treatment records, and the employment of advanced statistical methodologies all serve as pivotal pillars reinforcing the robustness and depth of our investigation. Collectively, these elements underscore the invaluable contributions of our study to the realm of ACS research. However, our study inherits several limitations: A major limitation of the ABC Study was that the diagnosis of myocardial infarction did not account for troponin measurement, as it was not in use at that time; therefore, we used the rise and gradual decline of creatine kinase and creatine kinase-MB as biochemical markers of necrosis, Nevertheless, these markers of necrosis are still recommended in the absence of troponin measurement [28]. Additionally, at the time of patient enrolment, percutaneous transluminal coronary angioplasty (PTCA) was not yet used to reopen coronary arteries in patients with STEMI, Thus, whether the results may have been altered by early mechanical reperfusion remains uncertain. However, our findings align with a recent study conducted by Hurskainen et al., involving 8,049 ACS patients, where over 85% of STEMI patients underwent primary percutaneous coronary intervention. Finally, since the patients in this study were exclusively of Caucasian descent. Therefore, caution must be exercised when implementing the current findings on other demographic groups and ethnic populations.

## CONCLUSIONS

The ABC study provided insights into the long-term incidence and risk factors for stroke among ACS patients. Notably, it identified a number of clinical predictors associated with a higher risk, including older age, AF, and a higher ACR. Conversely, a higher eGFR was correlated with a lower risk of stroke. Additionally, the study identified a geographical association with the risk of fatal stroke. This underscores the importance of considering both individual clinical predictors and broader geographic factors in stroke prevention policies.

## ACKNOWLEDGEMENTS

The authors thank Paola Michelazzo, RN; Jessica Civiero, RN; and nurses from the emergency care units of Conegliano, Adria, and Bassano General Hospitals for their assistance with patient management. We thank Renzo De Toni, PhD; Patrizio Buttazzi, PhD; Giancarlo Battistella, RN and the general laboratory personnel for assistance in collecting laboratory data. We thank Professor Paolo Palatini (Padova University) for his scientific and intellectual support from the beginning of the ABC study which continued for many years.

## DATA AVAILABILITY STATEMENT

The data underlying this article will be shared on reasonable request to the corresponding author.

